# When lockdown policies amplify social inequalities in COVID-19 infections. Evidence from a cross-sectional population-based survey in France

**DOI:** 10.1101/2020.10.07.20208595

**Authors:** Nathalie Bajos, Florence Jusot, Ariane Pailhé, Alexis Spire, Claude Martin, Laurence Meyer, Nathalie Lydié, Jeanna-Eve Franck, Marie Zins, Fabrice Carrat, for the SAPRIS study group

## Abstract

**Objectives:** To assess social inequalities in the trends in COVID-19 infections following lockdown

**Design:** A cross-sectional survey conducted among the general population in France in April 2020, during COVID-19 lockdown.

**Participants:** 10 401 participants aged 18-64, from a national cohort who lived in the three metropolitan French regions most affected by the first wave of COVID-19.

**Main outcome:** The main outcome was occurrence of possible COVID-19 symptoms, defined as the occurrence of sudden onset of cough, fever, dyspnea, ageusia and/or anosmia, that lasted more than three days in the 15 days before the survey. We used multinomial regression models to identify social and health factors related to possible COVID-19 before and during the lockdown.

**Results:** In all, 1,304 (13.0%; 95% CI: 12.0%-14.0%) reported cases of possible COVID-19. The effect of lockdown on the occurrence of possible COVID-19 was different across social hierarchies. The most privileged class individuals saw a significant decline in possible COVID-19 infections between the period prior to lockdown and during the lockdown (from 8.8% to 4.3%, P=0.0001) while the decline was less pronounced among working class individuals (6.9% before lockdown and 5.5% during lockdown, P=0.03). This differential effect of lockdown remained significant after adjusting for other factors including history of chronic disease. The odds of being contaminated during lockdown as opposed to the prior period increased by 57% among working class individuals (OR=1.57; 95% CI: 1.0-2.48). The same was true for those engaged in in-person professional activities during lockdown (OR=1.53; 95% CI: 1.03-2.29).

**Interpretation:** Lockdown was associated with social inequalities in the decline in COVID-19 infections, calling for the adoption of preventive policies to account for living and working conditions. Such adoptions are critical to reduce social inequalities related to COVID-19, as working-class individuals also have the highest COVID-19 related mortality, due to higher prevalence of comorbidities.

**Section 1: What is already known on this topic:** Significant differences in COVID-19 incidence by gender, class and race/ethnicity are recorded in many countries in the world. Lockdown measures implemented throughout the globe have been effective in reducing transmission risks.

**Section 2: What this study adds:** Our study shows that lockdown’s impact was socially differentiated and has benefited the working classes the least. Such results underline the need to design COVID-19 preventive policies that take into account living and working conditions, as working-class individuals also have the highest COVID-19 related mortality, due to higher prevalence of comorbidities.

## Introduction

Given the pre-existing social inequalities in health within societies^1^ and the significant differences in COVID-19 mortality by gender, class and origin recorded in countries such as France^2-3^, the United Kingdom^4^, the USA^5^ and other countries around the world^6^, several studies address issues of social inequalities related to COVID-19^7-11^. However, to our knowledge, no study has investigated potential social inequalities in the effects lockdown policies, widely implemented around the globe.

Our hypothesis is that lockdown measures, shown to be effective in reducing the number of new cases ^12^, have not been effective in the same way for all, failing to protect the most vulnerable populations. While more privileged social classes may have had greater exposure to the virus prior to lockdown, due to more frequent social interactions in public spaces (e.g. bars, restaurants) and travelling, they may have better adapted to lockdown measures, through telework, while working classes may have benefited less from lockdown conditions, due to their professional obligations as essential workers and their living conditions in overcrowded housing.

Our objective was to study the differential effect of lockdown measures on possible COVID-19 infections according to social class in France, one of the most affected countries in Europe by the first wave of COVID-19.

## Methods

### Study design and participants

The SAPRIS (*SAnté, Pratiques, Relations et Inégalités Sociales en population générale pendant la crise COVID-19*) survey was set-up mid-March 2020, with the general aim of understanding the main epidemiological, social and behavioural challenges of the SARS-CoV2 epidemic in France. It relies on a *consortium* of four prospective cohort studies involving three general population-based adult cohorts and a population-based children cohort. The analysis presented here is based on data from one of the three adult cohorts, the *Constances* cohort, which is the only cohort to have accurate data on professional status and preventive measures in the workplace. *Constances* is a generalist cohort made up of a national sample of 215 000 adults aged 18 to 69 at inclusion and recruited from 2012 onwards^14^.

All cohort members of *Constances* who had regular access to the internet (n=66,848) were invited to complete the SAPRIS questionnaire online. 69.0% participated in the survey (46,107). To best highlight the impact of the lockdown on possible COVID-19 symptoms, we chose to center this analysis on individuals (18-64 years) who have already been employed, living in one of the three metropolitan French regions most affected by the first wave of COVID-19 *i*.*e*. Grand Est, Ile-de-France (Paris Region) and Hauts-de-France. 10,101 participants met this criteria and were included in the analysis.

### Ethics and public involvement

The survey was approved by the National Institute for Health and Medical Research (Inserm) ethics evaluation committee (approval #20-672 dated March 30^th^, 2020).

### Data collection

Data collected online from April 6^th^ to May 5^th^, 2020 solicited information on socio-demographic characteristics, household size and composition, employment characteristics, daily life conditions, childcare arrangements, alcohol and tobacco use, sexual life, comorbidities, health care utilization and treatments. The questionnaire also addressed COVID-19 related topics including preventive behaviors (gel, mask, social distancing) for individuals and in the workplace, risk perceptions and COVID-19 related beliefs as well as a detailed description of COVID-19 symptoms over the last two weeks.

Symptoms were reported if they were unusual and occurred at least once in the past 15 days. The duration of symptoms were graded on a scale of one to five (less than 1 day, 1 to 3 days, 4 to 7 days, 8 to 14 days, >14 days). Finally, the total time (in days) between the onset of the first symptom and the date of the survey was reported.

### Measures

Our main outcome was a three-category measure, distinguishing 1) No suspicion of Covid-19 contamination, 2) probable contamination before the lockdown and 3) probable contamination during the lockdown. We used the following criteria defined by the European Centre for Disease Prevention to identify “possible COVID-19 contamination: at least cough or fever or dyspnea or sudden onset of ageusia, dysgeusia or anosmia occurring during the at-risk period”^15^. We added an additional criterion of duration, including symptoms lasting more than 3 days to add additional specificity to our definition.

The likely period of contamination (LPC) was identified as a function of *i)* the duration between the onset of the first symptoms and the date of the survey (DFS), *ii* the duration of incubation (DI: the 75th percentile duration between exposure and the onset of the first COVID-19 symptom) of 7 days^16^ and *iii)* the date of survey (DS). LPS was defined as follows:

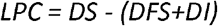

Based on this information probable contamination before the lockdown included LPC before March 17^t^h while contamination during the lockdown included LPC on or after March 17^t^h.

Participants’ social position was defined according to 3 criteria: current professional status (Inactive, retired or unemployed before the beginning of the pandemic/ employed but stopped working since the beginning of the pandemic, Full-time teleworking, Full-time or part-time in-person professional activities), socio-professional class and financial situation as perceived by respondents (comfortable/no problems/difficult). Socio-professional class was based on current or previous occupation, and distinguished health professions with specific exposure to the virus. The following 5 categories were constructed: Health professionals (doctors, nurses, caregivers), Upper class (senior managers), High middle class (intermediate professions), Low middle class (employees and skilled workers with a diploma of higher or equal to two years university degree), working class (unskilled employees and workers with a diploma lower than a two years university degree).

### Statistical methods

We used inverse probability weighting to correct for selection and non-participation biases. Weights were estimated using logistic regression models, with selection or participation as the response variables, and socio-demographics characteristics as covariates: sex, age group, occupational status (active, inactive), social affiliation and department of residence.

Since the information on the number of rooms in the housing unit was only asked in a second survey in June 2020, this information was missing for the 22% of the sample who didn’t complete the second questionnaire. We imputed this data using predictions obtained by logistic regression.

We first conducted bivariate analysis to explore the association between sociodemographic characteristics area (size of the agglomeration and region), number of individuals living in the household *per* room, educational level, nationality (French or not French), professional status, smoking, body mass index, health status (chronic diseases), and COVID-19 related behaviors (individual and workplace preventive measures (gel, mask, social distancing) and possible COVID-19 contamination in three categories.

We then conducted a multinomial logistic regression to compare the risk of contamination before (reference category) and during the lockdown according to social class, with successive and additional adjustments for other socio-demographic and health factors. The final model presents the variables that allow us to test our hypotheses on the effect of living conditions: housing, social class and professional status. We performed a sensitivity analysis including those with symptoms lasting less than 3 days and found results similar in magnitude but some became statistically non-significant (not shown).

All analyses were performed using R software. A P-value <0.05 was considered statistically significant. All percentages are weighted to account for the complex sampling design and post stratification.

Multivariable analyses were performed on unweighted data.

### Role of the funding source

The funders had no role in the design, analysis, interpretation or writing. All the authors had full access to all the data and NB and FC had final responsibility for the decision to submit for publication.

## Results

Table 1 describes the sociodemographic and health characteristics distribution of study participants and the frequency of possible COVID-19, according to the probable date of contamination.

**Table 1:** Participants characteristics and associated proportion of possible COVID-19 by period

|  | Weighted distribution | likely contamination | P-Value* | likely contamination prior the lockdown | likely contamination during the lockdown | P-Value** |
| --- | --- | --- | --- | --- | --- | --- |
| **Age** |  |  | 0.003 |  |  | 0.002 |
| 18-34 | 25.2 (1759) | 13.8 (232) |  | 8.3 (145) | 5.5 (87) |  |
| 35-44 | 29.7 (3028) | 14.8 (425) |  | 8.3 (267) | 6.6 (158) |  |
| 45-54 | 25.8 (2816) | 12.6 (378) |  | 8.7 (268) | 3.9 (110) |  |
| 55-64 | 19.3 (2498) | 9.7 (269) |  | 6.2 (193) | 3.5 (76) |  |
| **Sex** |  |  | 0.947 |  |  | 0.783 |
| Female | 52.4 (5164) | 13 (663) |  | 8.2 (442) | 4.8 (221) |  |
| Male | 47.6 (4937) | 13 (641) |  | 7.8 (431) | 5.2 (210) |  |
| **Nationality** |  |  | 0.437 |  |  | 0.709 |
| French | 96.1 (9703) | 13.1 (1263) |  | 8.1 (842) | 5.1 (421) |  |
| Not french | 3.9 (348) | 11.3 (37) |  | 7.3 (28) | 4 (9) |  |
| Missing | 50 |  |  |  |  |  |
| **Social Class** |  |  | 0.965 |  |  | 0.234 |
| Upper class | 32.7 (5541) | 13 (718) |  | 8.8 (500) | 4.3 (218) |  |
| Upper middle class | 20.9 (2397) | 12.9 (303) |  | 8.5 (205) | 4.4 (98) |  |
| Lower middle class | 18.1 (894) | 13.4 (114) |  | 8 (72) | 5.4 (42) |  |
| Working class | 21.8 (838) | 12.4 (108) |  | 6.9 (66) | 5.5 (42) |  |
| Health professional | 6.5 (431) | 13.9 (61) |  | 6.2 (30) | 7.7 (31) |  |
| **Professional status** |  |  | 0.018 |  |  | 0.002 |
| Did not work before lockdown | 24.5 (2178) | 10.6 (241) |  | 7.2 (177) | 3.5 (64) |  |
| Employed and stopped working since COVID | 18.0 (1185) | 15.7 (181) |  | 10.3 (116) | 5.4 (65) |  |
| Switched to teleworking | 38.8 (5469) | 12.9 (719) |  | 8.1 (492) | 4.7 (227) |  |
| Continued face-to-face working | 18.7 (1269) | 13.7 (163) |  | 6.6 (88) | 7.1 (75) |  |
| **Overcrowding** |  |  | 0.077 |  |  | 0.183 |
| less or equal than one pers/room | 94.4 (9557) | 12.8 (1210) |  | 7.9 (816) | 4.9 (394) |  |
| more than one pers/room | 5.6 (544) | 16.3 (94) |  | 9.5 (57) | 6.7 (37) |  |
| **Financial resources** |  |  | 0.046 |  |  | 0.077 |
| At ease | 29.5 (4082) | 12.8 (493) |  | 8.3 (346) | 4.5 (147) |  |
| No particular problem | 46.5 (4369) | 12.1 (563) |  | 7.5 (382) | 4.5 (181) |  |
| Difficult | 24.1 (1581) | 15.2 (238) |  | 8.7 (138) | 6.6 (100) |  |
| Missing | 69 |  |  |  |  |  |
| **Region** |  |  | 0.02 |  |  | 0.006 |
| Ile-de-France | 38.7 (5195) | 14.4 (714) |  | 8.9 (482) | 5.5 (232) |  |
| Grand Est | 30.6 (2854) | 13.4 (368) |  | 9.1 (257) | 4.3 (111) |  |
| Hauts-de-France | 30.7 (2052) | 10.9 (222) |  | 5.8 (134) | 5.1 (88) |  |
| **Agglomeration size** |  |  | 0.21 |  |  | 0.268 |
| Rural area | 6.9 (488) | 15.2 (67) |  | 10.1 (47) | 5.1 (20) |  |
| < 50 000 | 4.3 (335) | 12 (44) |  | 7.3 (30) | 4.7 (14) |  |
| 50-200 000 | 8.2 (536) | 11.6 (67) |  | 7.5 (49) | 4.1 (18) |  |
| 200-500 000 | 17.1 (1972) | 10.9 (229) |  | 7.6 (157) | 3.3 (72) |  |
| 500 000 -1 000 000 | 24.8 (1564) | 12.2 (180) |  | 6.5 (106) | 5.6 (74) |  |
| > 1 000 000 | 38.8 (5200) | 14.4 (715) |  | 8.9 (482) | 5.5 (233) |  |
| Missing | 6 |  |  |  |  |  |
| **Chronic disease** |  |  | <0.00001 |  |  | <0.00001 |
| None | 77.7 (7923) | 12.9 (1001) |  | 7.8 (664) | 5 (337) |  |
| Hypertension | 4.7 (455) | 10.2 (54) |  | 4.7 (33) | 5.5 (21) |  |
| Asthma or other respiratory diseases | 2.6 (246) | 28.8 (61) |  | 19.2 (44) | 9.6 (17) |  |
| Diabetes, cancer, heart disease, heart disease, immune diseases, liver, kidney, immunity, | 2.4 (257) | 12.6 (34) |  | 10 (24) | 2.6 (10) |  |
| Others | 12.7 (1220) | 11.8 (154) |  | 7.5 (108) | 4.3 (46) |  |
| **Active smoking** |  |  | 0.807 |  |  | 0.856 |
| Yes, daily | 11.7 (819) | 12.9 (93) |  | 7.3 (57) | 5.6 (36) |  |
| Yes, sometimes (less than once a day) | 4.6 (348) | 11.3 (43) |  | 6.2 (26) | 5.1 (17) |  |
| No | 83.8 (8727) | 13.1 (1136) |  | 8.1 (770) | 4.9 (366) |  |
| Missing | 207 |  |  |  |  |  |
| **Obesity** |  |  | 0.811 |  |  | 0.448 |
| BMI<30 | 85.9 (8889) | 12.9 (1133) |  | 7.8 (758) | 5.1 (375) |  |
| BMI>30 | 14.1 (868) | 13.3 (124) |  | 9.1 (84) | 4.1 (40) |  |
| Missing | 344 |  |  |  |  |  |
| **Individual preventive measures (mask, gel, social distancing) during outings in the last 7 days.** |  |  | <0.00001 |  |  | <0.00001 |
| All 3 | 27 (2797) | 16.2 (413) |  | 9.7 (277) | 6.5 (136) |  |
| At least one | 63 (6355) | 11.3 (750) |  | 7 (497) | 4.4 (253) |  |
| None | 10 (949) | 14.7 (141) |  | 10 (99) | 4.6 (42) |  |
| **Preventive measures at work (mask, gel, social distancing)** |  |  | 0.75 |  |  | 0.036 |
| All 3 | 12.2 (851) | 14.1 (113) |  | 7.5 (65) | 6.6 (48) |  |
| At least one | 9.4 (721) | 13.0 (83) |  | 5.8 (43) | 7.2 (40) |  |
| None | 78.4 (8529) | 12.8 (1108) |  | 8.4 (765) | 4.5 (343) |  |
| **All** | **100 (10,101)** | **13.0 (1304)** |  | **8.0 (873)** | **5.0 (431)** |  |

\* *Chi2 test likely contamination/no contamination*
\*\* *Chi2 test between no contamination/prior/during the lockdown*

The sample was equally divided between men (47.6%) and women (52.4%) and the mean age was 43.50 years (95%CI: 43.17-43.83). More than a third (38.8%) of the sample lived in cities with more than 1,000,000 inhabitants while a minority (6.9%) lived in rural areas. About a third of the sample (32.7%) were considered upper class while 21.8% were working class. 15.3% of the sample had in person professional activities during the lockdown period. Altogether, 13.0% (95% CI: 12.0%-14.0%) of participants reported symptoms compatible with possible cases of COVID-19 (n=1335) in the two weeks preceding the survey.

Residents from the Paris region (Ile-de-France) (P=0.02), participants facing financial difficulties (P=0.046) and those who reported chronic conditions (asthma or respiratory pathologies specifically) (P<0.0001) were more likely to report possible COVID-19 while older participants (P=0.003), and those who did not work before lockdown (P=0.033) were less likely to report those symptoms. Reporting possible COVID-19 was unrelated to social class.

While the percentage of participants reporting possible COVID-19 infection during lockdown was lower than participants reporting possible COVID-19 infection before lockdown (5.0% *versus* 8.0%), this decrease was uneven across social groups. As shown in Figure 1, the decline was most pronounced among privileged classes (from 8.8% before lockdown to 4.3% during lockdown, P=0.001) while the decline was least pronounced among the working class (from 6.9% before lockdown to 5.5% during lockdown, P=0.03).

**Figure 1:**
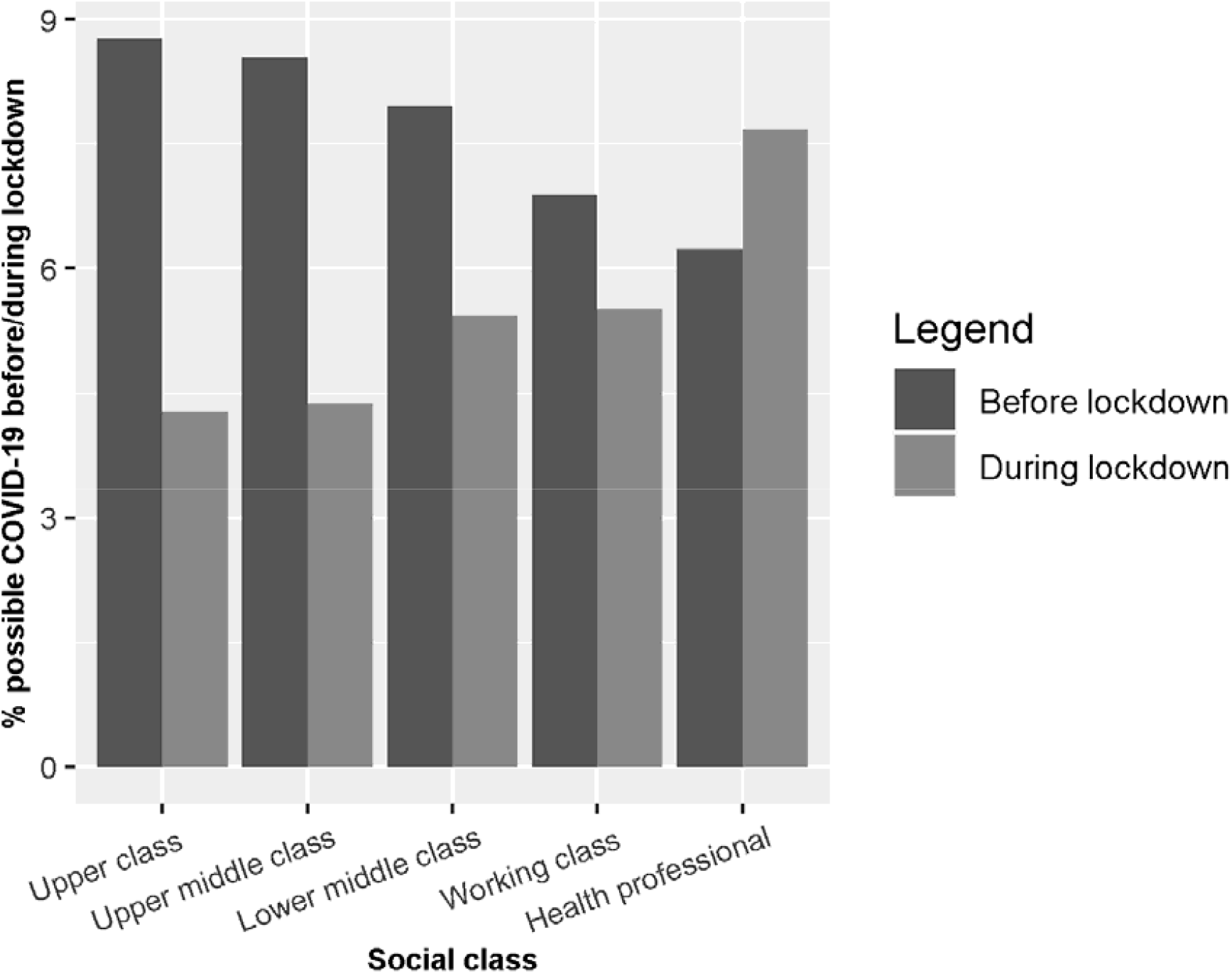
Percentage of individuals likely to be contaminated before or during lockdown by social class.

In addition, those living in housings with less than one room per person were slightly more likely to report a possible case of COVID-19 than others (16.3% versus 12.8%, P=0.08), with no difference between before and during lockdown.

The multivariable analyses presented in Table 2 indicated that the odds of no contamination relative to probable infection prior lockdown was unrelated to social class but depended on the region of residence, with increased odds among residents from the Hauts-de-France region relative to those residing in the Paris region (Ile de France) (OR= 1.39; 95% CI: 1.13-1.71).

**Table 2:**
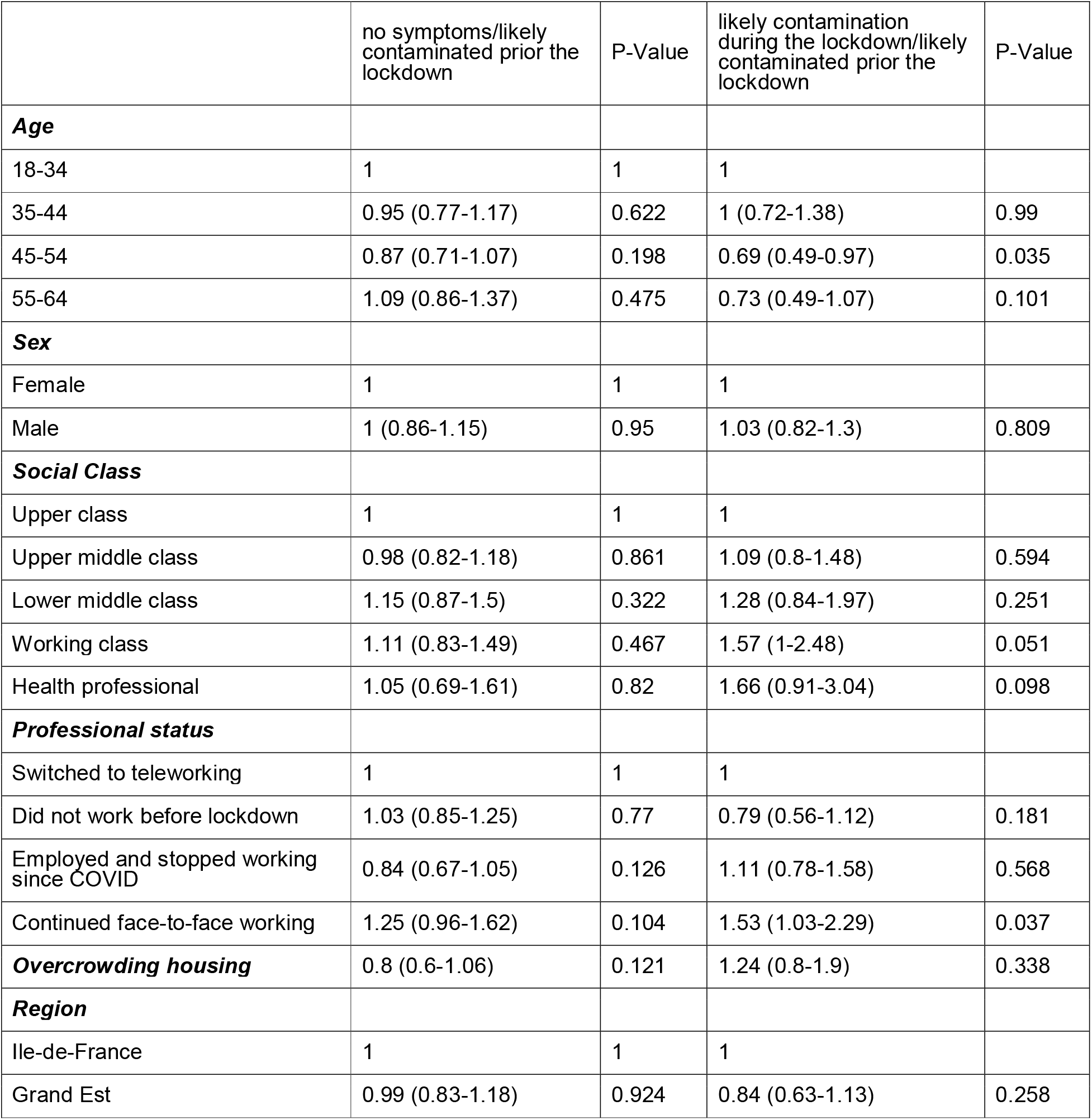
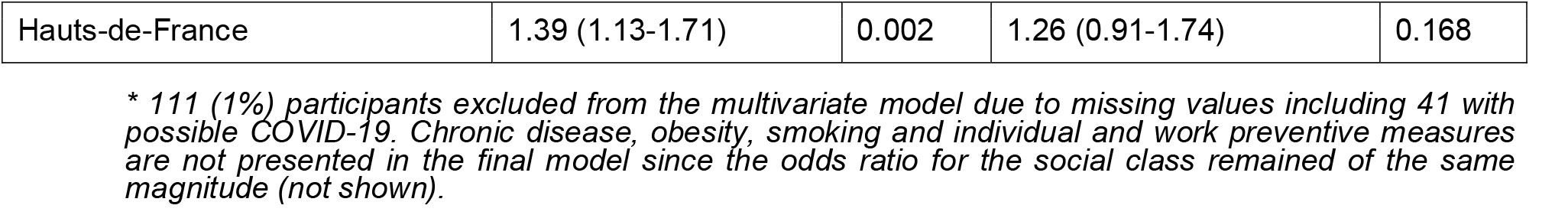
Factors associated with possible COVID-19: adjusted OR (95% CI) Multinomial regression results *Reference group: probable contamination prior to the lockdown* *OR adjusted for all the variables presented in the table*

Regarding the risk of infection during lockdown relative to the risk of infection before lockdown, it was higher among participants who had in-person professional activities compared to those who worked remotely (OR=1.53; 95% CI: 1.03-2.29). This risk was also increased among working class compared to upper class participants (OR=1.57; 95% CI: 1-2.48). It is worth noting that the odds-ratio for working class was 1.53 (95% CI: 0.96-2.42) when adjusting for smoking and it was 1.49 (95%CI: 0.93-2.4) when adjusting for history of chronic disease and obesity. Finally, this odds-ratio was reduced to 1.39 (95% CI: 0.87-2.21) when adjusting for perceived financial situation.

To study the stability of our results, we performed a sensitivity analysis check by repeating the multivariate analyses without excluding individuals who reported symptoms lasting less than 3 days. The odds ratio remained of the same magnitude but the degrees of significance were lower (not shown).

## Discussion

To our knowledge, SAPRIS is the largest general population-based COVID-19 study in Europe that simultaneously collects detailed data on symptoms and social characteristics to investigate the impact of lockdown on possible COVID-19 infections.

Analyses by time period, corresponding to whether individuals may have been infected before or during lockdown, show differential trends by social class that were masked in an overall analysis. The issue of temporality is essential because confinement measures affected individuals differently according to their housing and working conditions^5,17^. Individuals at the top of the social hierarchy saw a greater decline in COVID-19 symptoms after the lockdown than those from the working class. In fact, working-class individuals were more likely than those in the upper class to have been contaminated during lockdown rather than before.

Our results show that this overexposure during lockdown was partly a result of their health status (smoking and history of chronic disease and obesity). It was also partly an effect of their economic precariousness since the OR of the working class decreased when it was adjusted on this variable, a result consistent with economic work that has recently been established at a macroeconomic level in France^18^. We also found that living in housing with less than one room per person tended to be linked to the risk of having been contaminated. Finally, our results do not reflect a lower propensity of the working class to adopt individual prevention measures.

One can think that the overexposure to the virus of the working class during lockdown may reflect, at least in part, the fact that more individuals belonging to this class live in neighborhoods with high population density. Such an effect is not completely captured by the size of the agglomeration. For example, the density in some neighborhoods in the Paris suburbs, where excess mortality by COVID-19 is particularly high, is higher than that observed in larger cities^19^. Residents of these dense cities could have faced a higher risk of being exposed to the virus by encountering contagious individuals in shops, in the streets, or in public transports.

In any case, the data suggest that working class individuals were less protected by the lockdown measures than the more privileged categories.

This analysis has several limitations. First, the sample is socially diverse but is not fully representative of the French population as it only represents three regions in France and respondents from the Constances cohort who have internet connectivity are not representative of all residents in France. In particular, the study fails to capture particularly vulnerable groups such as undocumented migrants and homeless people, who are particularly affected by the pandemic^8^.

While the study provides information on social status based on education and employment, it doesn’t capture other forms of social disadvantage including race and ethnicity that are shown to increase the risk of COVID-19 infection in many settings and the risk COVID-19 related mortality in France^3^ and other countries^20-22^.

Additionally, it should be noted that our analyses are based on reported symptoms rather than on biologically tested cases, thus excluding asymptomatic individuals. However, the shortage of tests did not permit the use of testing in this study conducted in the early stages of the pandemic, especially before lockdown, as the use of RT-PCR testing was limited to patients with severe symptoms. Our symptom-based analysis is nevertheless consistent with epidemiological surveillance data by region^19^ and data on over-exposition of individuals with chronic respiratory diseases^23^.

Another limitation relates to the fact that some people may have had COVID-19 symptoms prior to the 15 days of the survey and are not counted in our possible COVID-19 cases. Since the socio-demographic structure of the respondents is stable during the study period (not shown), it is reasonable to think that the *de facto* exclusion of these situations does not affect results on association of possible Covid19 with social class.

In addition, although symptom reporting may risk being socially differentiated, it is reasonable to assume that any social reporting bias does not vary during the month of the survey.

In any case, from a prevention perspective, it is important to characterise the most exposed social groups and to try to uncover the social logics that favour this exposure, particularly those referring to living conditions^24,25^.

In conclusion, we showed that the effect of a lockdown policy designed and applied without taking into account social characteristics can contribute to increasing social inequalities in exposure to the risk of contracting the virus, as was rightly pointed out recently by Anderson et al.^26^ In this sense, the biomedical approach to prevention, which promotes preventive measures based on clinical knowledge without taking into account the socially differentiated effects of living conditions shows its limits, as was the case in the fight against previous epidemics^9,27^. Our results call for the implementation of future preventive policies that tackle these social inequalities. Such implementation is critical to reduce social inequalities related to COVID-19, as working-class individuals also have the highest COVID-19 related mortality, due to higher prevalence of comorbidities.

## Data Availability

Data are available upon reasonable request

## Acknowledgments

The authors warmly thank all the volunteers of the Constances cohort.

We thank the staff of the Constances cohort that have worked with dedication and engagement to collect and manage the data used for this study and to ensure continuing communication with the cohort participants.

We thank A Sireyjol and L Kuhn for their help in statistical analysis and Pr J Bouyer for his advice in multivariable analysis.

We thank Rosalind Bell-Aldeghi for her help in editing the manuscript.

## *The SAPRIS study group

The SAPRIS study group: Nathalie Bajos (co-Principal investigator), Fabrice Carrat (co-Principal investigator), Pierre-Yves Ancel, Marie-Aline Charles, Florence Jusot, Claude Martin, Laurence Meyer, Alexandra Rouquette, Ariane Pailhé, Gianluca Severi, Alexis Spire, Mathilde Touvier, Marie Zins.

## Data Statement

The datasets used and analyzed during the current study are available from the corresponding author on reasonable request.

## Funding

### This study

ANR (Agence Nationale de la Recherche, #ANR-20-COVI-000,#ANR-10-COHO-06), Inserm (Institut National de la Santé et de la Recherche Médicale, #C20-26).

### Cohort funding

The CONSTANCES Cohort Study is supported by the Caisse Nationale d’Assurance Maladie (CNAM), the French Ministry of Health, the Ministry of Research, the Institut national de la santé et de la recherche médicale. CONSTANCES benefits from a grant from the French National Research Agency [grant number ANR-11-INBS-0002] and is also partly funded by MSD, AstraZeneca, Lundbeck and L’Oreal.

## Notes

### Competing Interest Statement

The authors have declared no competing interest.

### Funding Statement

ANR (Agence Nationale de la Recherche, #ANR-20-COVI-000,#ANR-10-COHO-06), Inserm (Institut National de la Sante et de la Recherche Medicale, #C20-26).

### Author Declarations

The survey was approved by the National Institute for Health and Medical Research (Inserm) ethics evaluation committee (approval #20-672 dated March 30th, 2020).

